# Prevalence of Pathogenic Germline Variants in Cancer Susceptibility Genes using the All of Us Dataset

**DOI:** 10.1101/2025.03.28.25324849

**Authors:** Gideon Idumah, Daphne Newell, Madeleine Hadrys, Isabella Ribaudo, Ying Ni, Joshua Arbesman

**Author notes:** These authors contributed equally (Gideon Idumah, Daphne Newell). ***Correspondence:*** Joshua Arbesman, MD, Department of Dermatology, 9500 Euclid Avenue, Cleveland, OH 44195, USA.

## Abstract

**Importance:** Understanding the prevalence of the pathogenic germline variants in cancer susceptibility genes within a non-selected population could be highly beneficial for evaluating the need for broader genetic testing guidelines and thereby aiding early detection of cancer.

**Objective:** To determine the population-level prevalence of pathogenic variants in specific cancer susceptibility genes, which are typically screened for only in individuals with cancer or their at-risk relatives due to cost considerations. We will further breakdown the variant prevalence by race, ethnicity and sex

**Design:** Cancer susceptibility genes were selected based on the Invitae Multi-Cancer panel.

**Setting:** This population-based genomic study utilized short-read whole genome sequencing (srWGS) data from the All of Us (AoU) controlled tier database v8.

**Participants:** A total of 414,830 individuals with srWGS data and variant information were included in the analysis.

**Main Outcomes and Measures:** We extracted the ClinVar variant consequences for each participant and applied stringent criteria to classify variants as truly pathogenic or likely pathogenic (P/LP). A total of 3,454 unique P/LP variants were identified across 77 transcripts and 72 genes.

**Results:** We identified 20,968 individuals with P/LP variants, representing approximately 5.05% of the total population. Pathogenic variant prevalence did not significantly differ by sex and ethnicity, while significant differences were observed across racial groups (adj p < 0.0001), with white participants having the highest prevalence (5.72%) and Asians the lowest. Among individual genes, *MUTYH* (1.33%) had the highest prevalence, followed by *BRCA2* (0.42%) and *MITF* (0.37%). Pathogenic variants in *TP53* showed significant sex-based differences, while a higher number of genes had significantly different carrier rates across ethnicity (7) and racial groups (14).

**Conclusions and Relevance:** The pathogenic variant prevalence found in the general population, along with its racial variability, highlights the need to reconsider current genetic testing guidelines. Expanding screening recommendations could enhance early cancer detection and prevention efforts ultimately reducing disease burden and improving outcomes.

**Key Points:** *Questions:* What are the population-level prevalence rates of pathogenic germline variants in specific cancer susceptibility genes?

*Findings:* Among 414,830 participants with short read whole genome sequencing data from the All of Us dataset, 5.05% carried a pathogenic or likely pathogenic variant in one of 84 cancer susceptibility genes identified from the Invitae Multi-Cancer Panel.

*Meaning:* These findings suggest that pathogenic variants are sufficiently common to support broader genetic screening programs for cancer predisposition, enabling early detection and targeted prevention strategies.

## Introduction

Germline genetic testing for cancer susceptibility has typically been reserved for individuals with known risk factors, such as those with a personal history of cancer or a family member with a pathogenic variant.^1^ However, with increasing accessibility and affordability, broader genetic screening is becoming more feasible. Current National Comprehensive Cancer Network (NCCN) guidelines recommend genetic testing when the probability of detecting a high-penetrance breast susceptibility variant exceeds 5%, based on prior-probability models.^2^ Likewise, a simulation study for Lynch Syndrome found that a 5% risk threshold made population-based genetic screening both cost-effective and beneficial to health outcomes.^3^ If the prevalence of P/LP variants in the general population reaches this clinically actionable threshold, expanding genetic testing recommendations may be warranted. This expansion must balance clinical benefit, cost analysis and a healthcare system’s capacity to manage increased testing while considering the advantages of early cancer detection.

While prior studies have focused on the prevalence of P/LP variants among cancer patients, the prevalence in a general, diverse population has yet to be characterized.^4–6^ Our study utilized the All of Us (AoU) biobank, which links whole genome sequencing data to electronic health records (EHR), allowing for an assessment of P/LP variant prevalence across a diverse population representative of the U.S. We aim to provide insights that could shape future genomic medicine strategies for cancer screening.

## Methods

We analyzed the AoU research program’s-controlled tier dataset v8, available to authorized users on the researcher workbench, which includes genomic data from 414,830 individuals with short-read genome sequencing. The dataset includes 2,180,727 single nucleotide variants (SNVs) and indels, with multiallelic sites split into separate records.

To focus our analysis on cancer susceptibility genes (CSGs), we considered the 84 genes included in the Invitae Multi-Cancer Panel, which are associated with primarily adult-onset, hereditary cancers across major organ systems.^7^ To ensure the accuracy of the transcripts identified by the panel, we cross-checked with the NCBI ClinVar database, and only retained transcripts that were either classified as MANE or MANE Plus Clinical.^8^

Using the AoU variant annotation table, we employed the following criteria to identify P/LP variants in the selected transcripts. First, we removed any transcripts whose consequences is annotated as ‘downstream_gene_variant’ or ‘upstream_gene_variant’. In addition, we excluded all *MC1R* variants and all *EPCAM* variants except for deletions. Next, any variant labeled as P/LP but also included other ambiguous annotations such as risk factor, uncertain significance or likely allele were manually verified in the ClinVar website. After applying these criteria, we identified 3,454 unique P/LP variants across 77 transcripts and 72 genes.

## Results

We identified 20,968 individuals carrying P/LP variants, representing 5.05% of the cohort. Table 1 summarizes the prevalence of these individuals across sex, ethnicity and race. Statistical analyses using two-sided z-tests (for sex/ethnicity) and Chi-Square tests (for race) revealed no significant differences in sex and ethnicity, but a significant difference across racial groups (adj p < 0.0001). For gene-by-gene analysis, we applied the Bonferroni correction for multiple testing. White participants had the highest prevalence (5.72%), while Asian participants had the lowest.

**Table 1:** Demographic distribution of participants analyzed.

|  |  | Entire Cohort | CSG-Carrier |  | *p* value |
| --- | --- | --- | --- | --- | --- |
|  |  |  | N | (%) |  |
| **Sex** | Male | 159,163 | 7,944 | 4.99% | 1.326e-01 |
|  | Female | 247,637 | 12,622 | 5.10% |  |
| **Ethnicity** | Hispanic | 79,704 | 3,942 | 4.95% | 1.250e-01 |
|  | Non-Hispanic | 323,509 | 16,430 | 5.08% |  |
| **Race** | White | 223,997 | 12,809 | 5.72% | p<0.0001 |
|  | Asian | 13,490 | 439 | 3.25% |  |
|  | Black or African American | 72,538 | 2549 | 3.51% |  |
|  | American Indian or Alaska Native | 5,754 | 239 | 4.15% |  |
|  | Middle Eastern or North African | 2,351 | 95 | 4.04% |  |
|  | Native Hawaiian or Other Pacific Islander | 456 | 21 | 4.61% |  |

Gene-specific analyses (Figure 1), showed that *MUTYH* had the highest prevalence (1.33%), followed by *BRCA2* (0.42%) and *MITF* (0.37%). We further analyzed the results by sex, ethnicity and race (Figs 1B-D) to reveal: *TP53* was the only gene with a significant sex difference (adj p = 0.02), while seven genes had significant prevalence differences between Hispanics and non-Hispanics individuals. Racial differences were observed in 14 genes, with MUTYH showing the greatest variability-1.7% in White individuals, 1.4% in Middle Eastern individuals, and 0.4% in Asian individuals. Full data are available in the supplementary materials.

**Figure 1:**
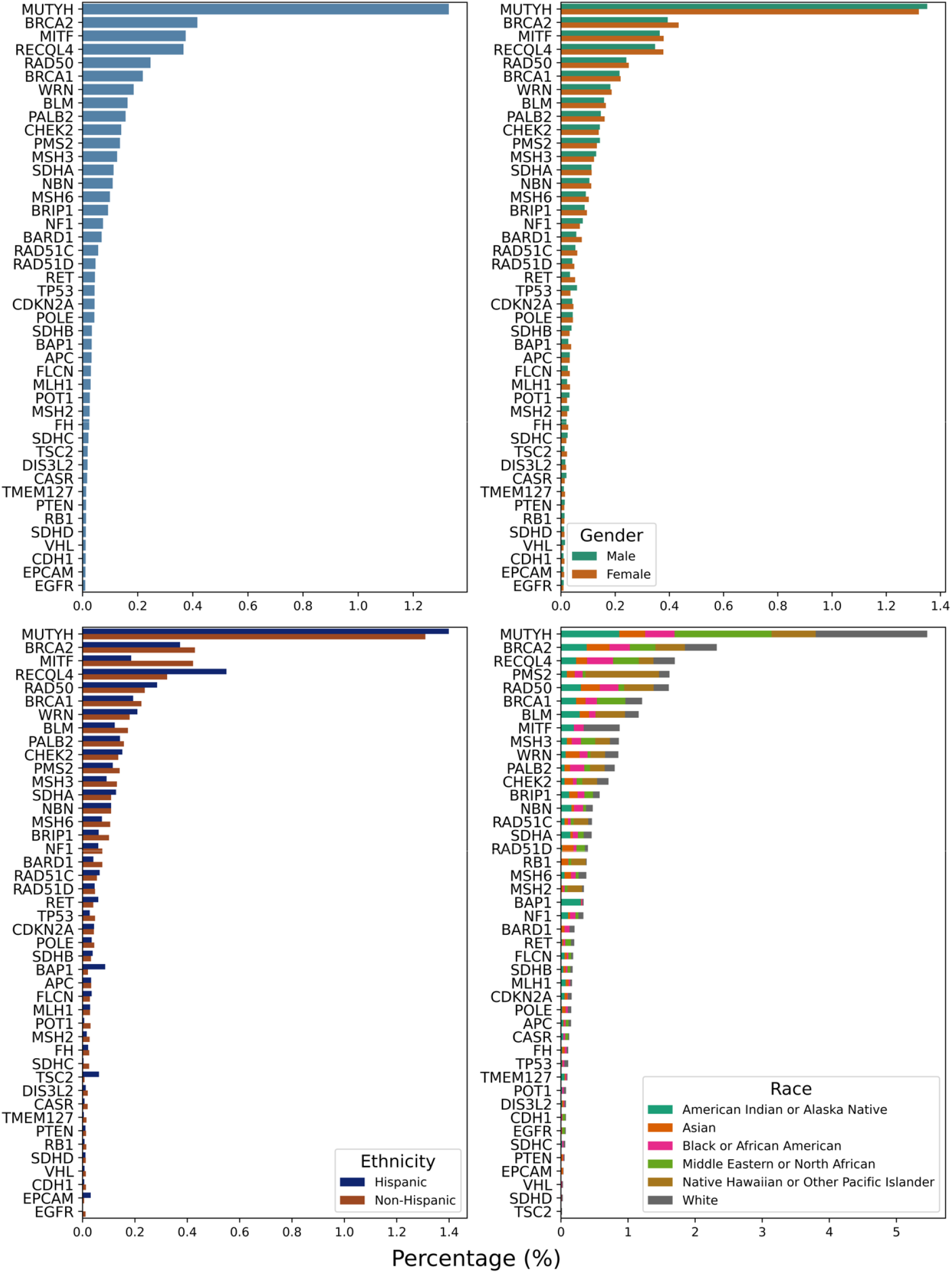
Gene-level distribution of individuals with P/LP variants. We have excluded genes with an insignificant number of affected individuals for clarity. Each individual was counted only once, even if they had multiple variants in multiple genes (A) Plot of the percentages of unique individuals with P/LP variants for each gene. B-D) Further breakdown of pathogenicity rates by sex, race, and ethnicity per gene.

## Discussion

Our findings suggest that the 5.05% general population prevalence of P/LP variants in cancer susceptibility genes aligns with the NCCN’s 5% threshold for genetic testing recommendations, supporting broader genetic screening initiatives. However, implementing a population-based screening for germline cancer susceptibility requires further cost-effectiveness analyses and evaluating whether the healthcare system could accommodate such changes. Despite these challenges, cancer screening in individuals with germline susceptibility has been proven to save lives and reduce costs.^9^

It is important to consider that this could lead to personalized preventive strategies and inform public health policies. In particular, the reported population frequency of pathogenic *BRCA1/2* mutations in non-Ashkenazi Jewish populations has been estimated to be 1 in 400 (0.25%) to 1 in 800 (0.13%). However, we found the prevalence of *BRCA1* and *BRCA2* carriers to be significantly higher at 0.22% and 0.42%, respectively, which is in line with an analysis of ExAC.^10^ Additionally, recent studies using AoU data suggest underdiagnosis of Lynch syndrome in the general population.^11^ Similarly, another recent analysis found the prevalence of thyroid cancer-associated syndrome genes in AoU to be ~10-20 times higher than currently estimated in the general population.^12^ Along with our findings, this indicates significantly higher rates of cancer susceptibility variants in an unselected population, and therefore we estimate that standard clinical practice and genetic testing guidelines miss a significant number of individuals who would benefit from cancer susceptibility identification via genetic testing.

There has been caution toward using genomic research to inform clinical practice due to the lack of ethnic and racial diversity in genomic studies.^13^ Many of these studies are conducted in populations with primarily European ancestry, and thus may bias clinical practice to benefit these populations.^14^ The AoU dataset is uniquely diverse with 77% of participants from historically underrepresented communities in biomedical research, and 46% identifying as racial and ethnic minorities.^15^ Our findings, therefore, offer valuable insights that could guide more inclusive genetic screening recommendations. Lastly, sensitivity to individual anxiety secondary to genetic testing will be a critical component of pre-test counseling as we consider test expansion to a non-cancer population.

## Limitations

This study has limitations. First, the AoU dataset relies on volunteer participants, introducing potential healthy volunteer bias. While this is a concern for disease penetrance studies, it is less relevant here as we focused on genetic variant prevalence. Second, demographic categorizations were based on self-identified race and ethnicity in EHRs, which may not fully capture the complexity of genetic ancestry. We caution against applying racial/ethnic prevalence differences directly to clinical decision-making but recognize their potential to refine genetic screening guidelines. Thus, we do not make specific recommendations for genetic testing based upon variable variant rates in particular racial and ethnic groups.

## Conclusions

Our study provides the first large-scale analysis of cancer susceptibility P/LP variant prevalence in a diverse, unselected population. Using whole genome sequencing data from the AoU dataset we identified 5% of individuals carrying P/LP variants, a prevalence aligning with the NCCN’s 5% genetic testing threshold. While no significant differences were observed across sex and ethnicity, notable racial disparities highlight the need for more inclusive genetic research. Future studies should further assess the cost-effectiveness, clinical utility, and feasibility of expanded genetic testing in diverse populations. Addressing these challenges will help advance medicine towards a more proactive and equitable approach to cancer prevention, ultimately reducing disease burden and improving public health outcomes.

## Data Availability

All data produced in the present study are available upon reasonable request to the authors

## Acknowledgements

We gratefully acknowledge *All of Us* participants for their contributions, without whom this research would not have been possible. We also thank the National Institutes of Health’s *All of Us* Research Program for making available the participant data examined in this study.

## Author Contributions

*Concept and design*: Idumah, Newell, Ni, Arbesman.

*Acquisition, analysis, or interpretation of data:* Idumah, Newell, Hadrys

*Drafting of the manuscript:* Idumah, Newell, Ribaudo, Ni, Arbesman

*Critical revision of the manuscript for important intellectual content:* All authors

*Statistical analysis:* Idumah.

*Supervision:* Ni, Arbesman.

